# IL-32 in limb-girdle muscular dystrophy LGMDR1-calpain 3 related: A Pilot Study on Its Role as a Biomarker

**DOI:** 10.64898/2025.12.18.25342268

**Authors:** Jenita Immanuel, Andrea Valls, Cristina Ruiz, Juan José Poza, Patricia Garay-Albizuri, Roberto Fernández-Torrón, Adolfo López de Munain, Amets Sáenz

## Abstract

In LGMDR1-Calpain-3 related, as in all muscular dystrophies, clinical trial monitoring remains a challenge due to the lack of reliable biomarkers. This study assessed IL-32 concentrations in both serum and urine, uncovering a marked increase in patients compared to healthy controls. Serum IL-32 levels were especially elevated in young adults, suggesting a possible link to the early and more active phases of disease onset. Meanwhile, urinary IL-32 levels showed consistent elevation across all age groups, reinforcing its promise as a stable, non-invasive biomarker. These findings support the potential of IL-32 in monitoring disease progression and therapeutic response in clinical trials, and underscore its potential involvement in LGMDR1.

## Introduction

Limb-girdle muscular dystrophy type R1-calpain 3 related (LGMDR1) or Calpainopathy, the most widespread form of LGMD is an autosomal recessive disease characterized by progressive atrophy of the proximal limb muscles and shoulder girdle muscles^1,2^. The known cause of this disease are the mutations in calpain-3 gene, coding a calcium-activated protease ^3^, which lead to muscle damage significantly impacting mobility and quality of life ^2,4–6^. Patients often experience difficulty walking, climbing stairs, or lifting objects with symptoms often beginning in adolescence and gradually progressing to require mobility aids such as wheelchairs^1,2^.

LGMDR1 research faces key challenges including genetic and clinical heterogeneity, slow disease progression, and a lack of standardized outcome measures. These factors complicate diagnosis, trial design, and outcome evaluation of therapeutic trials. There’s an urgent need for reliable biomarkers to enable early detection, monitor disease activity, and assess treatment response ^7,8^.

Inflammation is one of the main cause of pathophysiology of diseases implicated in skeletal muscle dysfunction^9^. Krahn and colleagues (2011) described several patients initially diagnosed with eosinophilic myositis (EM) who were later found to have mutations in the *CAPN3* gene, confirming a diagnosis of LGMDR1. These patients showed elevated serum creatine kinase (CK), muscle weakness and eosinophilic infiltration in early muscle biopsies ^10,11^. Notably, eosinophils were more common in younger patients and absent in older ones, suggesting that eosinophilic myositis may be an early and transient feature of calpainopathy ^12^. Additionally, our group reported elevated IL-32 expression in LGMDR1 muscle biopsies with eosinophil infiltration, highlighting its potential role in modulating immune responses in muscular disorders ^13^.

IL-32 is a pro-inflammatory cytokine involved in autoimmune, infectious, cancerous, and skin disorders. It promotes the production of other cytokines and modulates immune cell behavior, contributing to chronic tissue inflammation ^14^. In recent decades, many researches have been conducted to demonstrate the wide-range of functions of IL-32 ^14–16^.

Building on previous findings that associates LGMDR1 with eosinophilic infiltration and immunological activation, for the first time, we have examined IL-32 concentration in LGMDR1 patient’s serum and urine samples and compared to healthy controls. These findings will support IL-32 as a practical and accessible biomarker, with strong potential for use in future clinical studies focused on diagnosis, disease monitoring, and therapeutic targeting in muscular dystrophies.

## Methods

Human samples from age-matched healthy controls and LGMDR1 patients were supplied by the “Donostia University Hospital, Spain” following each participant’s voluntary signing of an informed consent form before sample collection. Enzyme-linked immunosorbent assay (ELISA) was employed to quantify IL-32 levels in both serum and urine samples. ELISA assay was first conducted in serum samples, a well-established platform for biomarker evaluation, to enable a more comprehensive characterization. To complement these findings, IL-32 was also measured in urine, for its advantages of non-invasive collection, broader sample availability, and suitability for longitudinal monitoring.

In serum samples, the mean age (average ± SD) of LGMDR1 patients (n = 7) and healthy controls (n = 8) were 33.25 ± 8.61 (ranging between 23 and 50) and 32.87 ± 10.85 (ranging between 20 and 60) years old, respectively. In urine samples, the mean age (average ± SD) of LGMDR1 patients (n = 18) and healthy controls (n = 13) of were 32.14 ± 15.33 (ranging between 13 and 62) and 33.0 ± 17.54 (ranging between 14 and 61) years old, respectively. According to North Star Assessment for LGMD (NSAD) and Vignos scale, the dystrophy’s functional stage was graded ^17,18^. Patient’s mutations and clinical information are detailed in Table I & II. The Human IL-32 (Interleukin 32) ELISA kit Assay system (EK242189, AFG scientific) was used to test serum/urine IL-32 in accordance with the manufacturer’s instructions. Using the dilution buffer included in the assay kit, the serum/urine samples from LGMDR1 patients and healthy volunteers were diluted 1:4 and 1:2, respectively, to bring the concentration of serum/urine IL-32 within the quantifiable range of the assay kit (15.63–1000 pg/mL).

**Table I.** Analyzed serum samples.

| Sample Number | Status | Mutation 1 | Mutation 2 | Age at Extraction | IL32 pg/ml | Vignos Scale |
| --- | --- | --- | --- | --- | --- | --- |
| SU-01 | LGMDR1 | Del. Calpain3 | X822LeuExt61X | 30-40 | 177,7 | 3 |
| 06-071 | LGMDR1 | Del. Calpain3 | X822LeuExt61X | <29 | 188,95 | 3 |
| 06-072 | LGMDR1 | Del. Calpain3 | X822LeuExt61X | <29 | 84,38 | 3 |
| 07-497 | LGMDR1 | 60dela | delEx5-7 | <29 | 215,49 | 4 |
| 06-164 | LGMDR1 | Del. Calpain3 | X822LeuExt61X | <29 | 100,84 | 9 |
| 02-011 | LGMDR1 | p.Arg788Serfs*14 | p.Arg788Serfs*14 | 30-40 | 97,12 | 9 |
| 97-0021 | LGMDR1 | p.Arg788Serfs*14 | p.Arg788Serfs*14 | >40 | 80,51 | 10 |
| 78-500-720 | Control | - | - | <29 | 42,33 |  |
| 78-500-719 | Control | - | - | <29 | 65,91 |  |
| 78-500-722 | Control | - | - | 30-40 | 43,47 |  |
| 78-500-723 | Control | - | - | >40 | 45,94 |  |
| 78-500-718 | Control | - | - | <29 | 44,92 |  |
| 78-500-737 | Control | - | - | 30-40 | 46,34 |  |
| A001 | Control | - | - | <29 | 41,88 |  |
| A002 | Control | - | - | >40 | 39,49 |  |

**Table II.** Analyzed Urine samples.

| Sample Number | Status | Mutation 1 | Mutation 2 | Age at Extraction | IL32 pg/ml | NSAD |
| --- | --- | --- | --- | --- | --- | --- |
| OR-01 (A) | LGMDR1 | p.(Arg788Serfs*14) | p.(Arg788Serfs*14) | <20 | 101,11 | 31/54 |
| OR-02 (A) | LGMDR1 | p.(Arg788Serfs*14) | p.(Arg788Serfs*14) | <20 | 160,93 | 29/54 |
| OR-03 (A)* | LGMDR1 | X822Leu ext61X | Del. Calpain3 | 31-45 | 101,46 | 3/54 |
| OR-04 (A) | LGMDR1 | p.(Arg788Serfs*14) | p.(Arg788Serfs*14) | 21-30 | 94,5 | 3/54 |
| OR-07 (A) | LGMDR1 | p.(Arg788Serfs*14) | p.(Arg788Serfs*14) | <20 | 105,65 | 31/54 |
| OR-18 (A) | LGMDR1 | p.(Arg788Serfs*14) | p.(Arg788Serfs*14) | <20 | 258,51 | 30/54 |
| OR-21 (A) | LGMDR1 | p.(Arg788Serfs*14) | 1782+1072 G>C | >46 | 141,3 | 3/54 |
| OR-25 (A) | LGMDR1 | p.(Arg489Trp) | p.(Arg788Serfs*14) | >46 | 88,62 | 3/54 |
| OR-26 (A)* | LGMDR1 | p.(Arg489Trp) | p.(Arg788Serfs*14) | 31-45 | 96,82 | 54/54 |
| OR-27 (A)* | LGMDR1 | P.(Arg748Gln) | p.(Arg788Serfs*14) | 31-45 | 119,22 | 9/54 |
| OR-31 (A) | LGMDR1 | N/A | N/A | >46 | 236,91 | N/A |
| OR-33 (A) | LGMDR1 | N/A | N/A | 21-30 | 137,83 | N/A |
| OR-42 (A)* | LGMDR1 | N/A | N/A | 21-30 | 139,11 | N/A |
| OR-46 (A) | LGMDR1 | N/A | N/A | 31-45 | 191,84 | N/A |
| OR-47 (A) | LGMDR1 | N/A | N/A | >46 | 118,19 | N/A |
| OR-48 (A) | LGMDR1 | N/A | N/A | >46 | 197,24 | N/A |
| OR-19 (B) | LGMDR1 | p.(Arg748Gln)) | p.(Arg788Serfs*14) | <20 | 120,88 | 54/54 |
| OR-29 (B) | LGMDR1 | p.(Gly222Arg) | p.(Arg101Lys) | 21-30 | 109,31 | 30/54 |
| OR-05 (A) | Control | - | - | >46 | 48,04 |  |
| OR-08 (A) | Control | - | - | 21-30 | 48,66 |  |
| OR-09 (A) | Control | - | - | 21-30 | 61,78 |  |
| OR-10 (A) | Control | - | - | 31-45 | 55,71 |  |
| OR-11 (A) | Control | - | - | <20 | 62,5 |  |
| OR-12 (A) | Control | - | - | <20 | 70,69 |  |
| OR-14 (A) | Control | - | - | <20 | 72,93 |  |
| OR-15 (A) | Control | - | - | >46 | 46,89 |  |
| OR-16 (A) | Control | - | - | <20 | 47,68 |  |
| OR-23 (A) | Control | - | - | >46 | 77,9 |  |
| OR-24 (A) | Control | - | - | >46 | 58,14 |  |
| OR-39 (A) | Control | - | - | <20 | 53,13 |  |
| OR-40 (A) | Control | - | - | <20 | 42,65 |  |

The statistical significance of the IL-32 concentrations in the control and patient groups was evaluated using an unpaired T-test. Using the Pearson correlation analysis, the correlation between IL-32 concentration and age or functional scale in LGMDR1 patients were assessed.

## Results

Quantitative analysis of IL-32 concentrations in both serum and urine revealed significant elevation in LGMDR1 patients compared to healthy controls. Serum IL-32 levels, were significantly elevated in the patient group, with a mean ± SD of 135.00 ± 56.78 pg/mL, compared to 46.29 ± 8.20 pg/mL in healthy controls (Figure IA). Urinary IL-32 concentrations were similarly increased in LGMDR1 patients, with a mean ± SD of 139.99 ± 50.16 pg/mL, versus 57.44 ± 11.14 pg/mL in healthy controls (Figure IB). Both comparisons were statistically significant (p ≤ 0.0001), reinforcing IL-32’s potential as a robust indicator of disease-associated inflammation.

**Figure I.**
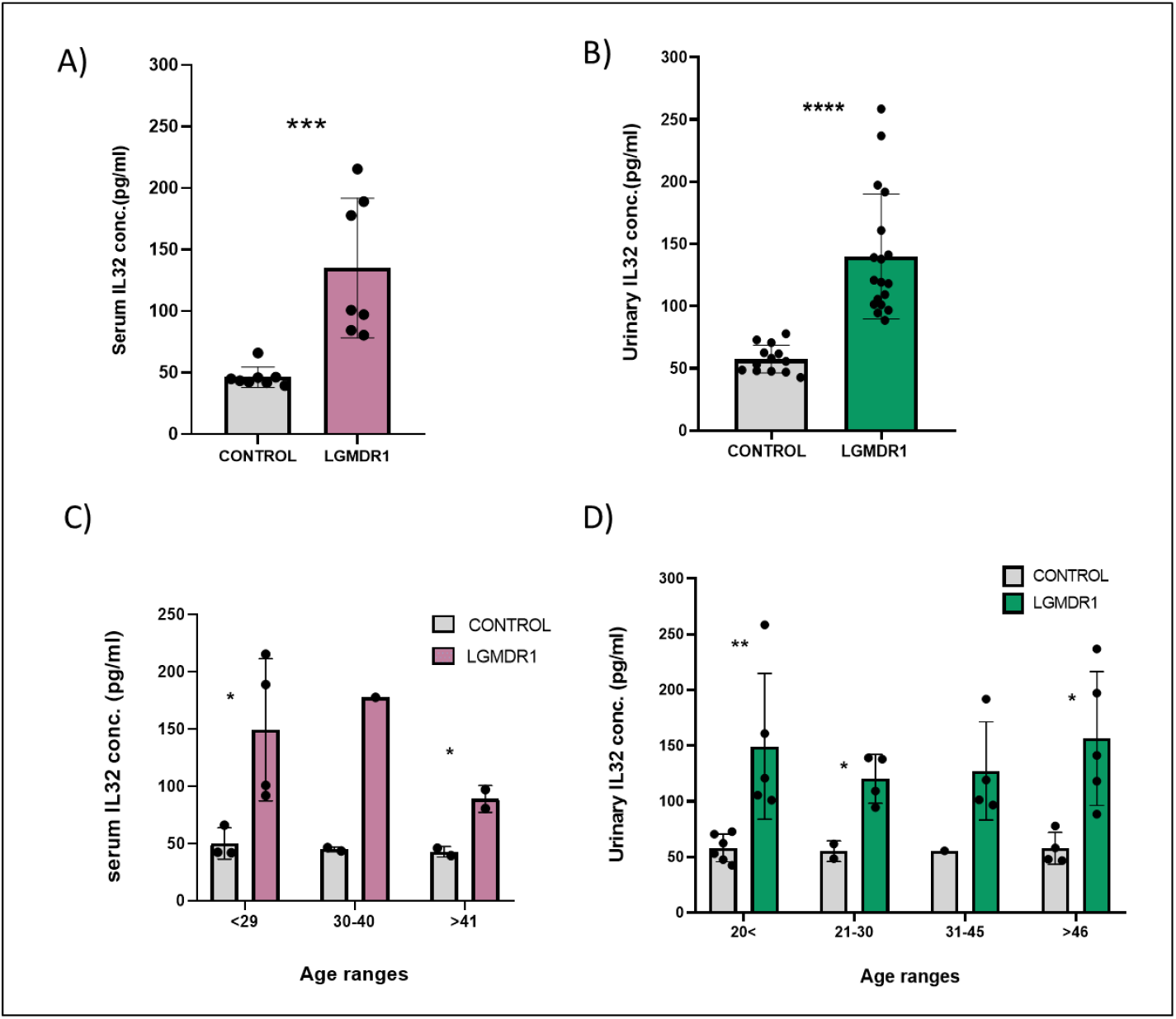
Serum and urinary IL-32 concentrations of healthy controls and LGMDR1 patients. (A) Serum IL-32 level of controls and patients. (B) Urinary IL-32 level of controls and patients. (C) Samples grouped by age for serum analysis. (D) Samples grouped by age for urine analysis. Error bars represent Standard Deviation (SD). *p ≤ 0.05; **p ≤ 0.01; ***p ≤ 0.001; ****p ≤ 0.0001

Given the wide inter-individual variability observed particularly among LGMDR1 patients, samples were stratified by age to better characterize IL-32 expression dynamics. Across all age groups, IL-32 concentrations remained elevated in patients compared to healthy controls in both serum and urine. Interestingly, in serum analysis, patients in early adulthood (less than 40 years of age) significantly exhibited the highest IL-32 levels, while it declined in individuals over 45, highlighting a more intense inflammatory response during the early phase of disease (p ≤ 0.01) (Figure 1C). In urine samples, IL-32 levels remained significantly elevated across all patient age groups compared to controls (p ≤ 0.05), reinforcing its potential as a consistent marker of inflammation (Figure ID).

Correlation analysis revealed a modest age-related decline in serum IL-32 levels among LGMDR1 patients (R^2^ = 0.250), with higher concentrations observed in early adulthood; healthy controls showed no correlation (R^2^ = 0.075) (Figure IIA). Urinary IL-32 levels remained consistently elevated across all patient age groups but showed no age-related trend (R^2^ = 0.012) (Figure IIB). Serum IL-32 also correlated strongly with phenotype severity based on the Vignos scale (R^2^ = 0.8987) (Figure IIC), whereas urinary IL-32 showed no association with functional status assessed by NSAD (R^2^ = 0.012) (Figure IID).

**Figure II.**
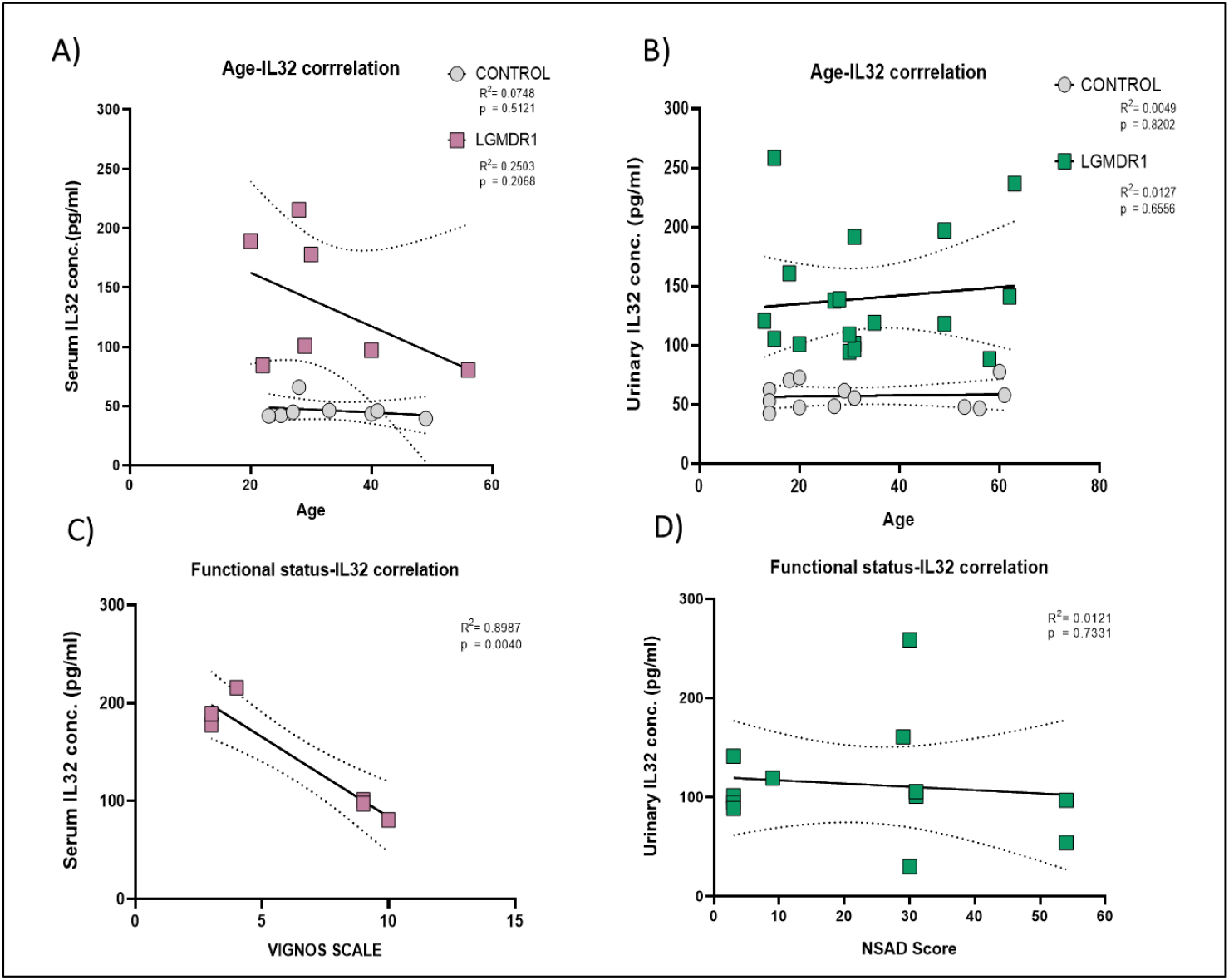
Correlation of IL-32 concentration and age or functional scale in serum and urine samples of healthy control and patients. (A) Age - IL-32 correlation in serum samples. (B) Age - IL-32 correlation in urine samples. (C) Functional status - IL-32 correlation in serum samples with Vignos scale. (D) Functional status - IL-32 correlation in urine samples with NSAD score.

## Discussion

Eosinophilic infiltration has also been observed in other muscular dystrophies such as dysferlinopathies and Becker muscular dystrophy ^19–21^. Among the inflammatory mediators implicated, IL-32 has emerged as a promising candidate since its upregulation was described in LGMDR1 pseudo-asymptomatic patients ^13^. For the best of our knowledge, IL-32 is not increased in these muscular dystrophies, but other interleukins, IL-27 and IL-17D, that showed increased expression, could be responsible of the inflammation in DMD and FSHD muscles respectively ^22,23^.

IL-32 is a pro-inflammatory cytokine capable of inducing TNF-α and IL-8, activating NF-κB and p38 MAPK pathways, and interacting with proteinase 3 (PR3) ^24^, potentially contributing to eosinophil recruitment. It has been also reported that IL-32 can act synergistically with NOD-mediated intracellular pathways to activate eosinophils ^25^. Its upregulation in LGMDR1 patients suggests a role in the immune response and positions IL-32 as a molecular bridge between muscle inflammation and genetic pathology ^13^. Its expression by both immune and nonimmune cells including fibroblasts and hepatocytes suggests a broader role in disease pathology ^2,9,27,28,10–16,26^. Given the involvement of CD56^−^ myofibroblasts in LGMDR1 ^8^, targeted investigation of IL-32 in these cells could clarify its role in disease pathology. IL-32 is typically induced by inflammatory stimuli and cellular stress, and its regulation by oxygen-sensing systems highlights its relevance to oxidative stress ^24^, a known contributor to LGMDR1 progression ^29^.

On the other hand, other studies have attempted to elucidate the functional significance of IL-32 binding to integrins. Moreover, the *in vitro* interaction between IL-32 and the extracellular domain of αVβ3 integrins has been reported ^24^. As in LGMDR1 integrin β1D mislocalization has been reported ^8^, further deep analysis are required to establish if a relationship between these two proteins exists.

Our comprehensive analysis of IL-32 levels in both urine and serum samples highlights its consistent elevation in LGMDR1 patients. The finding paves the way for understanding the inflammatory landscape and enhances its clinical utility: a) serum levels reflect systemic immune activity and a good correlation with the age/functional status of the patients and b) urine sampling, even without age/functional status correlation, allows for easy, repeatable monitoring over time.

Contrary to the observations for serum CK levels, IL32 levels remained elevated in wheelchair-bound LGMDR1 patients. Following the report by Krahn and colleagues (2006) of LGMDR1 patients with idiopathic EM^10^, subsequent investigations revealed eosinophilic infiltrations even in adulthood ^10,28,30^. Moreover, IL-32 was overexpressed in LGMDR1 patients muscles (1.81 fold change). However this was not considered a significant increase, since it did not exceed the two-fold threshold required to be considered substantially enhanced (Saenz et al. 2008, data not shown). IL32 measure is highly valuable for clinical trials, as muscle recovery testing is very limited in wheelchair-bound patients.

This dual detectability, combined with IL-32’s potential role in recruiting immune cells and activating inflammatory pathways, positions it as a biologically relevant and practical biomarker for LGMDR1, tracking disease progression, and evaluating therapeutic response.

## Data Availability

All data produced in the present work are contained in the manuscript

## Acknowledgments

The authors would like to thank the patients, their families and the healthy controls.

## Statements and declarations

### Datasets/Data Availability Statement

The data supporting the findings of this study are available within the article and/or its supplementary material.

## References

1. Fardeau, M. et al. Juvenile limb-girdle muscular dystrophy. Clinical, histopathological and genetic data from a small community living in the Reunion Island. Brain 119 (Pt 1, 295–308 (1996).

2. Urtasun, M. et al. Limb-girdle muscular dystrophy in Guipúzcoa (Basque Country, Spain). Brain 121 (Pt 9, 1735–1747 (1998).

3. Richard, I. et al. Mutations in the proteolytic enzyme calpain 3 cause limb-girdle muscular dystrophy type 2A. Cell 81, 27–40 (1995).

4. Ono, Y. et al. Functional defects of a muscle-specific calpain, p94, caused by mutations associated with limb-girdle muscular dystrophy type 2A. J. Biol. Chem. 273, 17073–17078 (1998).

5. Fanin, M. et al. Loss of Calpain-3 Autocatalytic Activity in LGMD2A Patients with Normal Protein Expression. Am. J. Pathol. 163, 1929–1936 (2003).

6. Lo, H. P. et al. Limb-girdle muscular dystrophy: diagnostic evaluation, frequency and clues to pathogenesis. Neuromuscul. Disord. 18, 34–44 (2008).

7. Petek, L. M. et al. A cross sectional study of two independent cohorts identifies serum biomarkers for facioscapulohumeral muscular dystrophy (FSHD). Neuromuscul. Disord. 26, 405–413 (2016).

8. Valls, A. et al. Urinary N-terminal titin fragment ascertained as biomarker in a small cohort of limb-girdle muscular dystrophy LGMDR1-calpain 3 related. J. Neuromuscul. Dis. 22143602251323628 (2025) doi:10.1177/22143602251323629.

9. Londhe, P. & Guttridge, D. C. Inflammation induced loss of skeletal muscle. Bone 80, 131–142 (2015).

10. Krahn, M. et al. CAPN3 mutations in patients with idiopathic eosinophilic myositis. Ann. Neurol. 59, 905–911 (2006).

11. Krahn, M. et al. Eosinophilic infiltration related to CAPN3 mutations: a pathophysiological component of primary calpainopathy? Clinical genetics vol. 80 398–402 at 10.1111/j.1399-0004.2010.01620.x (2011).

12. Wang, G. et al. Development of differential diagnostic models for distinguishing between limb-girdle muscular dystrophy and idiopathic inflammatory myopathy. Arthritis Res. Ther. 26, 215 (2024).

13. Sáenz, A. et al. Gene expression profiling in limb-girdle muscular dystrophy 2A. PLoS One 3, e3750 (2008).

14. Wallimann, A. & Schenk, M. IL-32 as a potential biomarker and therapeutic target in skin inflammation. Front. Immunol. 14, 1264236 (2023).

15. Kim, S. Interleukin-32 in inflammatory autoimmune diseases. Immune Netw. 14, 123–127 (2014).

16. Xin, T., Chen, M., Duan, L., Xu, Y. & Gao, P. Interleukin-32: its role in asthma and potential as a therapeutic agent. Respir. Res. 19, 124 (2018).

17. James, M. K. et al. Validation of the North Star Assessment for Limb-Girdle Type Muscular Dystrophies. Phys. Ther. 102, (2022).

18. Vignos, P. J. J., Spencer, G. E. J. & Archibald, K. C. Management of progressive muscular dystrophy in childhood. JAMA 184, 89–96 (1963).

19. Gallardo, E. et al. Inflammation in dysferlin myopathy: immunohistochemical characterization of 13 patients. Neurology 57, 2136–2138 (2001).

20. Weinstock, A., Green, C., Cohen, B. H. & Prayson, R. A. Becker muscular dystrophy presenting as eosinophilic inflammatory myopathy in an infant. J. Child Neurol. 12, 146–147 (1997).

21. Arahata, K. et al. Inflammatory response in facioscapulohumeral muscular dystrophy (FSHD): immunocytochemical and genetic analyses. Muscle Nerve. Suppl. 2, S56–66 (1995).

22. Haslett, J. N. et al. Gene expression comparison of biopsies from Duchenne muscular dystrophy (DMD) and normal skeletal muscle. Proc. Natl. Acad. Sci. U. S. A. 99, 15000–15005 (2002).

23. Osborne, R. J., Welle, S., Venance, S. L., Thornton, C. A. & Tawil, R. Expression profile of FSHD supports a link between retinal vasculopathy and muscular dystrophy. Neurology 68, 569–577 (2007).

24. Aass, K. R., Kastnes, M. H. & Standal, T. Molecular interactions and functions of IL-32. J. Leukoc. Biol. 109, 143–159 (2021).

25. Wong, C.-K., Dong, J. & Lam, C. W.-K. Molecular mechanisms regulating the synergism between IL-32γ and NOD for the activation of eosinophils. J. Leukoc. Biol. 95, 631–642 (2014).

26. Doody, A. et al. Defining Clinical Endpoints in Limb Girdle Muscular Dystrophy: A GRASP-LGMD study. Res. Sq. (2023) doi:10.21203/rs.3.rs-3370395/v1.

27. Tidball, J. G. & Wehling-Henricks, M. Damage and inflammation in muscular dystrophy: potential implications and relationships with autoimmune myositis. Curr. Opin. Rheumatol. 17, 707–713 (2005).

28. Oflazer, P. S., Gundesli, H., Zorludemir, S., Sabuncu, T. & Dincer, P. Eosinophilic myositis in calpainopathy: Could immunosuppression of the eosinophilic myositis alter the early natural course of the dystrophic disease? Neuromuscul. Disord. 19, 261–263 (2009).

29. Rajakumar, D., Alexander, M. & Oommen, A. Oxidative stress, NF-κB and the ubiquitin proteasomal pathway in the pathology of calpainopathy. Neurochem. Res. 38, 2009–2018 (2013).

30. Amato, A. A. ADULTS WITH EOSINOPHILIC MYOSITIS AND CALPAIN-3 MUTATIONS. Neurology 70, 730–731 (2008).

